# Association between perinatal mental health and reproductive and neonatal complications: a retrospective birth cohort study

**DOI:** 10.1101/2022.06.11.22276276

**Authors:** Jennifer D. Runkle, Kendra Risley, Manan Roy, Maggie M. Sugg

**Affiliations:** North Carolina Institute for Climate Studies, North Carolina State University, 151 Patton Avenue, Asheville, North Carolina, USA 28801; Department of Nutrition and Health Care Management, Appalachian State University, Boone, North Carolina, USA 28608; Department of Geography and Planning, Appalachian State University, P.O. Box 32066, Boone, North Carolina, USA 28608

**Keywords:** perinatal mood and anxiety disorders, serious mental illness, depression/anxiety, pregnancy, neonatal, maternal morbidity

## Abstract

**Background:** Maternal mental health as an important precursor to reproductive and neonatal complications remains understudied in the US, particularly in the Southeastern region, despite high medical costs and maternal morbidity, and infant burden.

**Objective:** This study sought to estimate the incidence of perinatal mental health disorders and the associated increased risk of leading pregnancy and infant complications.

**Methods:** A population-based retrospective birth cohort of delivery hospitalizations and readmissions was constructed for birthing populations in South Carolina, 1999 to 2017. Prevalence rates were calculated for perinatal mood and anxiety disorders (PMAD), severe mental illness (SMI), and maternal mental disorders of pregnancy (MDP). Poisson regression models using generalized estimating equations were used to estimate adjusted relative risks for the association between mental health conditions and severe maternal morbidity (SMM), hypertensive disorders of pregnancy (HDP), gestational diabetes (GD), cesarean delivery (CD), preterm birth (PTB), and low-birth weight (LBW).

**Results:** The most prevalent maternal mental condition was MDP (3.9%), followed by PMAD (2.7%) and SMI (0.13%). PMAD was associated with a higher risk of SMM, HDP, and CD, as well as a higher risk of PTB and LBW infants. SMI was associated with LBW, HDP, and CD. Pregnant populations with MDP were more at risk for SMM, PTB, HDP, LBW, and CD. Each maternal mental health outcome was associated with an elevated risk for hospital readmissions up to 45 days after delivery.

**Discussion:** Results demonstrated the escalating burden of PMAD and MDP for pregnant populations over time with important consequences related to maternal and infant morbidity.

## INTRODUCTION

One understudied research priority pertains to the influence of maternal mental health as an important precursor to maternal and infant morbidity (1-3). Mental health disorders, particularly anxiety and depression, are among the most common morbidities in perinatal women, affecting up to 20% of women during and after pregnancy (3, 4). In the United Kingdom, psychiatric issues are the leading cause of death for women in the first 12 months postpartum (3, 4). Mental health disorders are drastically under-diagnosed and under-treated in perinatal women, with just 30% of perinatal women with depression identified in a clinical setting (5, 6). For those 30% identified, less than 7% receive adequate treatment (5). Despite a high morbidity and mortality rate, perinatal mental health disorders remain understudied. Researchers have highlighted the need for more population-based epidemiological studies on the prevalence of these conditions during pregnancy and in the postpartum period (7).

The identification and treatment of mental health issues in pregnant women are crucial in preventing adverse maternal and neonatal outcomes. For pregnant women, poor mental health can increase the likelihood of preterm labor, postpartum mental health issues, preeclampsia, hypertension, diabetes, suicide, and infanticide (8-10). Infants born to women with untreated mental health disorders are at higher risk for low birth weight and long-term childhood and adolescent behavioral and neurodevelopmental problems (8-11). Additionally, untreated maternal mental health conditions cost the United States healthcare system upwards of $15 billion dollars every year (8). Untreated mental health issues in women from conception through 5 years postpartum cost the state of Texas alone over $2.2 billion in 2019 (11). In addition to the societal cost of untreated maternal mental health conditions, which often result in reduced productivity, greater use of public sector services such as SNAP benefits and Medicaid, involve higher long-term healthcare costs for the mother and the child (11).

Perinatal mental health surveillance remains understudied in the U.S., particularly in the Southeastern region, despite high-costs and a high maternal morbidity and infant burden. This study sought to estimate the incidence of perinatal mental health disorders and the associated increased risk of leading maternal and infant complications using a retrospective cohort in South Carolina, 1999-2017. A secondary aim involved examining the risk of postpartum readmissions for maternal mental health outcomes and estimating the excess cost of deliveries for women with a mental health condition. Findings will address an important knowledge gap on the psychological well-being of pregnant women and shed light on priority mental health conditions that need more attention during the critical period of pregnancy.

## METHODS

### Hospital delivery discharges and readmission data

A retrospective birth cohort design was used to longitudinally examine the maternal mental health experience for pregnant populations. Hospital delivery discharge hospitalizations were obtained from the SC Revenue and Fiscal Affairs Office Health and Demographics division for all hospital labor and deliveries in South Carolina from 1999 to 2017. Postpartum readmissions for up to 45 days after the delivery event were also obtained. Hospital delivery discharge and readmission visits were linked with infant birth and infant death records using a unique maternal identification number. Only delivery hospitalizations for women with a residence in South Carolina, with a gestation week of 20 weeks or more, and age 18-55 years were included in the final sample. The North Carolina State University’s Institutional Review Board deemed this research exempt.

### Mental health outcomes

Borrowing from McKee et al.(12), primary outcomes of interest were defined using the delivery record at discharge: 1) perinatal mood and anxiety disorders (PMAD), 2) serious mental illness conditions (SMI), and 3) maternal mental disorders of pregnancy (MDP). MDP is a diagnosis used to document any mental disorder complicating pregnancy, childbirth, or the puerperium (13). PMAD and SMI are specific diagnosis codes to record mood or anxiety disorders and severe mental illness during pregnancy and up to one year after delivery. For this analysis, we operationalize perinatal mental health to include 1) all mental disorders that preexist or recurred during gestation and incident conditions that emerged during the prenatal or postpartum periods (i.e., PMAD, SMI) and 2) any mental disorder that first emerged during pregnancy (i.e., MDP) (14).

All International Classification of Disease (ICD) diagnosis and procedure codes, Ninth Revision, Clinical Modification (ICD-9-CM), used to define each condition, can be found in Supplemental Table 1. In October 2015, the Centers for Medicare & Medicaid Services (CMS) and commercial health insurers transitioned from coding hospital administration records using ICD-9 to the International Classification of Disease (ICD) diagnosis and procedure codes, Tenth Revision, Clinical Modification (ICD-10-CM). We relied on published equivalence mapping to convert ICD-10 data to ICD-9 codes to maintain continuity in how conditions were coded throughout the study (15).

### Maternal and infant complications

Recent research has made the linkage between maternal mental health during the preconception, prenatal, and postpartum periods and increased risk for leading maternal obstetric complications, including severe maternal morbidity (SMM) (16, 17), preterm birth (PTB) (18), cesarean delivery (CD), either elective or emergent, hypertensive disorders of pregnancy (HDP) and gestational diabetes mellitus (GDM) (14). Pregnant populations with GDM are two-fold more likely to develop postpartum depression (19). Prenatal antidepressant usage has been associated with preeclampsia, although a recent systematic review revealed inadequate evidence of harm posed (20). For this analysis, maternal complications included: SMM, excluding blood transfusions, GDM, and CD (defined in Supp Table 1). A widely available algorithm published by the Centers for Disease Control and Prevention was used to identify 21 indicators for SMM while accounting for the length of stay, in-hospital maternal deaths, and delivery type (e.g., primary or repeat cesarean); excluding ectopic or molar pregnancy or deliveries in which abortive procedures were performed (21). A blood transfusion-related SMM diagnosis was omitted due to the incomplete record-keeping of hospitals concerning blood volume transferred. Infant complications included the following: LBW (i.e., defined as an infant weight at birth 2500 grams or less) (22) and PTB (i.e., defined as a birth before 37 completed weeks of gestation) (23).

### Covariates

Risk factors for a maternal mental disorders include maternal age younger than 20 years or older than 35 years, maternal infections, episodic or inadequate prenatal care, or other forms of maternal distress (e.g., nutritional deficiency or three or more pregnancies) (24). Covariates included the following maternal factors: age in years (19 or less, 20 to 29, 30 to 39, 40 or more), educational attainment (high school or less, some college, college, and more than college), race/ethnicity (non-Hispanic Black, Hispanic, Other, non-Hispanic White), insurance status (uninsured, Medicare/Medicaid, commercial insurance, U.S. Veteran Affairs (VA)), Kotelchuk prenatal care adequacy index (inadequate, intermediate, adequate, and adequate plus) (25), smoking status (yes/no), substance abuse (Supp Table 1)(26), infections (ICD-9-CM: 760.2), obesity (ICD-9-CM: 278), parity (no prior deliveries, one prior delivery, two or more prior deliveries) and year of discharge. Nonmetro compared to a metro maternal residence was coded using 2010 rural-urban community area codes (RUCA) to zip code areas by relying on the 5-tiered coding schema (27).

### Hospital cost data

We also compared the total charges (i.e., dollar amount) as received on the UB-92 billing file for each delivery hospitalization across maternal mental health conditions to estimate the excess cost for these conditions over time.

### Statistical analysis

Maternal characteristics were compared across each primary maternal mental health outcome using chi-square goodness of fit for categorical variables. To account for the correlated binary mental health outcomes captured longitudinally for each woman (28), we used a modified Poisson regression approach using robust error variances to estimate the group-specific incidence rate ratio per 10,000 deliveries (IRR) and the relative risk (RR) with 95% confidence intervals (95%CI) were computed for each bivariate association (29). The first set of models examined each primary mental health outcome (i.e., PMAD, SMI, MDP) separately adjusted for maternal risk factors. The second set of models examined individual primary mental health outcomes and the addition of maternal or infant health complications separately. The third set of models examined the interaction between maternal/infant complications and maternal race/ethnicity. The fourth set of models examined the risk of postpartum hospital readmissions for maternal mental health outcomes separately. All final models included covariate adjustment for maternal age, race/ethnicity, education, insurance payor, substance abuse, adequacy of prenatal care, parity, smoking, obesity, infections, and urban vs. rural residence.

We performed three separate sensitivity analyses. First, we examined PMAD disaggregated into depression and anxiety disorders. Next, we looked at the influence of preexisting mental health conditions. Finally, we examined the influence of racial segregation and poverty on the incidence of maternal mental disorders.

All statistical analyses were performed in SAS software (version 9.3; SAS Institute Inc, Cary, NC).

## RESULTS

### Characteristics of maternal mental health disorders

There was a total of 913,330 hospital delivery discharges, 24,201 (2.65%) of which involved PMAD, 1,230 (0.13%) for SMI, and 35,435 (3.88%) for MDP (Table 1, Table S2). The rate of PMAD and MDP has been steadily on the rise, increasing each year from 1999 to 2017 (Figure 1). The prevalence of PMAD was higher in white pregnant populations, persons with some college, and among those with adequate followed by inadequate prenatal care. The prevalence of SMI or MDP was much more likely among women who received inadequate prenatal care. Shared maternal risk factors for PMAD, SMI, and MDP included older maternal age (i.e., 30 to 39 years), Medicaid beneficiaries, low educational attainment, substance abuse during pregnancy, urban residence, obesity, presence of an infection, and those with at least one prior pregnancy (Table 1).

**Table 1.** Maternal characteristics across PMAD, SMI, and MDP mental health outcomes for the retrospective birth cohort, South Carolina 1999-2017.

|  | PMAD (n=24,201) |  |  |  | SMI (n=1,230) |  |  |  | MDP (n=35,435) |  |  |  |
| --- | --- | --- | --- | --- | --- | --- | --- | --- | --- | --- | --- | --- |
|  | IRR | RR | 95%CI |  | IRR | RR | 95%CI |  | IRR | RR | 95%CI |  |
| *Age group* |  |  |  |  |  |  |  |  |  |  |  |  |
| 19 years or less | 147.18 | 0.59 | 0.56 | 0.62 | 12.45 | 0.93 | 0.77 | 1.12 | 314.64 | 0.83 | 0.80 | 0.86 |
| 20-29 years | 343.80 | 1.38 | 1.34 | 1.42 | 13.51 | 1.01 | 0.89 | 1.15 | 418.18 | 1.10 | 1.07 | 1.12 |
| 30-39 years | 406.57 | 1.63 | 1.51 | 1.76 | 20.91 | 1.56 | 1.10 | 2.20 | 524.29 | 1.38 | 1.29 | 1.47 |
| 40+ years | 249.11 | 1.00 | ref |  | 13.41 | 1.00 | ref |  | 381.07 | 1.00 | ref |  |
| *Educational* |  |  |  |  |  |  |  |  |  |  |  |  |
| High School or Less | 225.33 | 0.80 | 0.77 | 0.84 | 18.79 | 4.18 | 3.20 | 5.47 | 446.73 | 1.68 | 1.62 | 1.74 |
| Some College | 324.88 | 1.16 | 1.11 | 1.20 | 12.11 | 2.70 | 2.03 | 3.57 | 388.55 | 1.46 | 1.41 | 1.52 |
| College | 274.95 | 0.98 | 0.93 | 1.04 | 3.40 | 0.76 | 0.47 | 1.23 | 249.32 | 0.94 | 0.88 | 1.00 |
| More Than College | 280.57 | 1.00 | ref |  | 4.49 | 1.00 | ref |  | 265.56 | 1.00 | ref |  |
| *Race/ethnicity* |  |  |  |  |  |  |  |  |  |  |  |  |
| non-Hispanic Black | 125.46 | 0.34 | 0.33 | 0.35 | 15.19 | 1.10 | 0.97 | 1.25 | 238.79 | 0.47 | 0.46 | 0.48 |
| Hispanic | 123.06 | 0.33 | 0.30 | 0.36 | 5.66 | 0.41 | 0.27 | 0.62 | 152.43 | 0.30 | 0.28 | 0.32 |
| Other | 134.47 | 0.36 | 0.34 | 0.39 | 6.75 | 0.49 | 0.35 | 0.69 | 159.99 | 0.31 | 0.29 | 0.34 |
| non-Hispanic White | 370.66 | 1.00 | ref |  | 13.82 | 1.00 | ref |  | 509.90 | 1.00 | ref |  |
| *Insurance Payor* |  |  |  |  |  |  |  |  |  |  |  |  |
| No insurance | 196.64 | 0.73 | 0.67 | 0.80 | 7.87 | 1.48 | 0.97 | 2.26 | 327.42 | 1.09 | 1.02 | 1.16 |
| Medicare/Medicaid | 271.30 | 1.01 | 0.98 | 1.04 | 21.00 | 3.96 | 3.39 | 4.62 | 476.90 | 1.59 | 1.55 | 1.62 |
| Private Insurance | 266.09 | 0.99 | 0.93 | 1.06 | 13.29 | 2.50 | 1.84 | 3.41 | 282.42 | 0.94 | 0.88 | 1.00 |
| VA | 268.38 | 1.00 | ref |  | 5.31 | 1.00 | ref |  | 300.82 | 1.00 | ref |  |
| *Prenatal Care* |  |  |  |  |  |  |  |  |  |  |  |  |
| Inadequate | 249.87 | 1.11 | 1.07 | 1.16 | 21.55 | 2.27 | 1.93 | 2.66 | 475.58 | 1.53 | 1.48 | 1.58 |
| Intermediate | 199.84 | 0.89 | 0.84 | 0.94 | 11.58 | 1.22 | 0.96 | 1.55 | 323.35 | 1.04 | 0.99 | 1.09 |
| Adequate | 318.32 | 1.42 | 1.38 | 1.46 | 13.25 | 1.39 | 1.21 | 1.61 | 416.37 | 1.34 | 1.31 | 1.37 |
| Adequate Plus | 224.26 | 1.00 | ref |  | 9.51 | 1.00 | ref |  | 311.00 | 1.00 | ref |  |
| *Substance Abuse* |  |  |  |  |  |  |  |  |  |  |  |  |
| Yes | 1112.36 | 4.31 | 4.06 | 4.58 | 126.28 | 10.42 | 8.51 | 12.76 | 5642.00 | 17.17 | 16.80 | 17.55 |
| No | 257.55 | 1.00 | ref |  | 12.12 | 1.00 | ref |  | 328.51 | 1.00 | ref |  |
| *Urban-rural* |  |  |  |  |  |  |  |  |  |  |  |  |
| Urban | 281.65 | 1.37 | 1.32 | 1.42 | 13.79 | 1.14 | 0.98 | 1.34 | 402.64 | 1.27 | 1.23 | 1.31 |
| Rural | 205.61 | 1.00 | ref |  | 12.12 | 1.00 | ref |  | 316.57 | 1.00 | ref |  |
| *Parity* |  |  |  |  |  |  |  |  |  |  |  |  |
| 0 | 273.86 | 1.16 | 1.13 | 1.20 | 12.02 | 0.98 | 0.86 | 1.12 | 338.24 | 0.71 | 0.69 | 0.73 |
| 1 | 319.22 | 1.36 | 1.32 | 1.40 | 17.55 | 1.43 | 1.25 | 1.64 | 384.18 | 0.81 | 0.79 | 0.83 |
| 2+ | 235.26 | 1.00 | ref |  | 12.24 | 1.00 | ref |  | 476.42 | 1.00 | ref |  |
| *Maternal infections* |  |  |  |  |  |  |  |  |  |  |  |  |
| Yes | 366.03 | 1.37 | 1.23 | 1.54 | 29.53 | 2.22 | 1.50 | 3.29 | 599.99 | 1.56 | 1.43 | 1.70 |
| No | 266.41 | 1.00 | ref |  | 13.31 | 1.00 | ref |  | 384.42 | 1.00 | ref |  |
| *Obesity* |  |  |  |  |  |  |  |  |  |  |  |  |
| Yes | 539.47 | 2.15 | 2.07 | 2.24 | 20.63 | 1.58 | 1.29 | 1.94 | 639.24 | 1.72 | 1.66 | 1.78 |
| No | 251.04 | 1.00 | ref |  | 13.03 | 1.00 | ref |  | 371.28 | 1.00 | ref |  |

PMAD= perinatal mood and anxiety disorders, SMI=severe mental illness, and MDP= mental disorders of pregnancy.
IRR = Incidence Rate Ratio, RR= Relative risk,
CI=Confidence Interval

**Figure 1.**
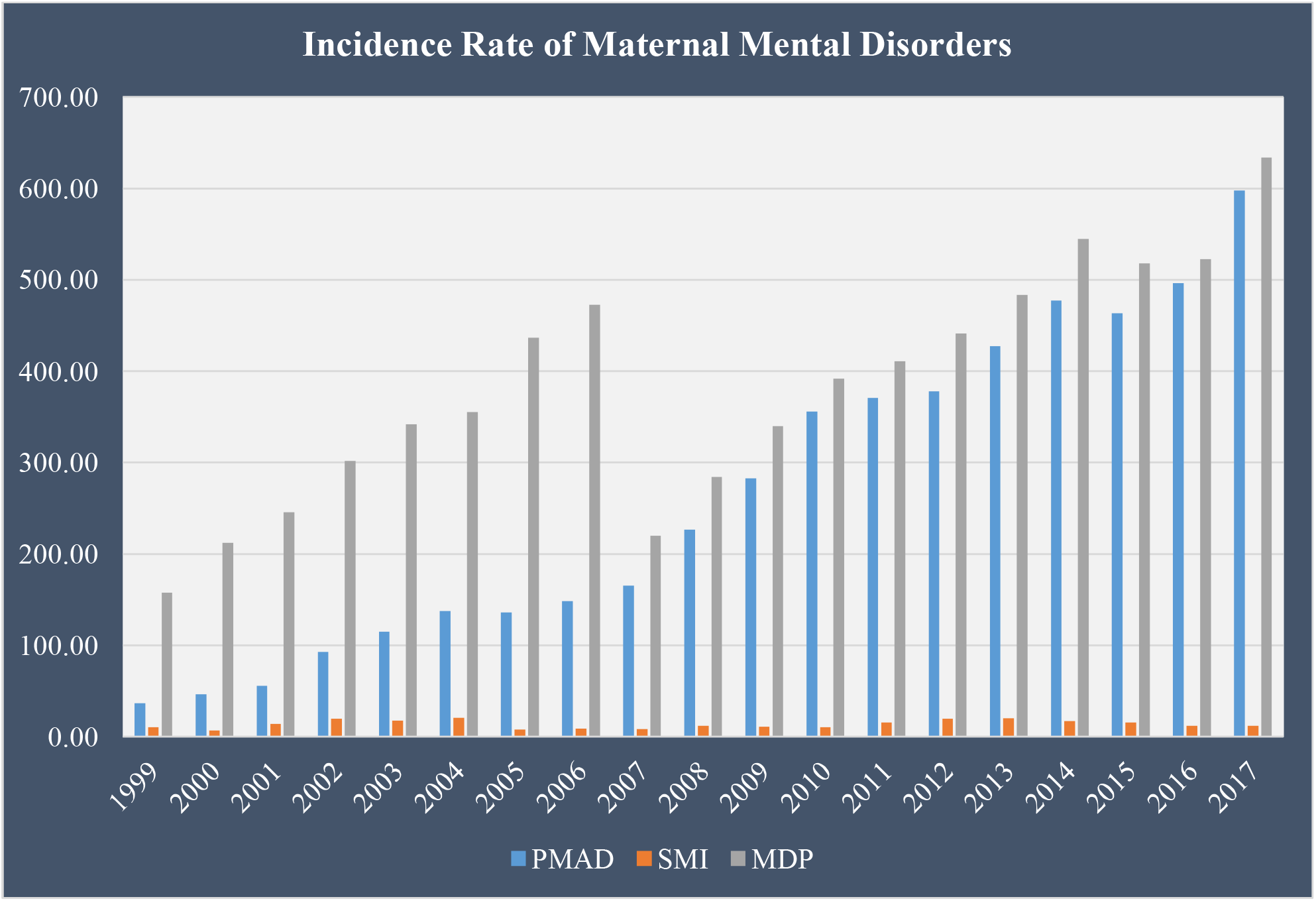
Annual incidence rates for Perinatal Mood and Anxiety Disorders (PMAD), Severe Mental Illness (SMI), and Mental Disorders of Pregnancy (MDP), South Carolina 1999-2017.

### Racial differences in PMAD, SMI, and MDP

After adjusting for other maternal factors, White compared to Black, Hispanic, or ‘other race’ pregnant populations were the most likely to be diagnosed with PMAD, SMI, and MDP (Table 2). However, compared to Hispanic birthing people, Black birthing people were 2.76 times (95%CI: 1.77, 4.30) and 1.43 times (95%CI: 1.32, 1.33) more likely to be diagnosed with SMI or MDP, respectively. We noted a similar elevated risk profile for SMI and MDP among women identifying as Black compared to those identifying as ‘other race’.

**Table 2.** Racial and ethnic differences in maternal risk of PMAD, SMI, or MDP, South Carolina delivery discharges 1999-2017.

| *Racial and ethnic differences* | PMAD |  |  | SMI |  |  | MDP |  |  | Anxiety |  |  | Depression |  |  |
| --- | --- | --- | --- | --- | --- | --- | --- | --- | --- | --- | --- | --- | --- | --- | --- |
|  | aRR | 95%CI |  | aRR | 95%CI |  | aRR | 95%CI |  | aRR | 95%CI |  | aRR | 95%CI |  |
| Black vs. Hispanic | 1.11 | 1.00 | 1.22 | 2.76 | 1.77 | 4.30 | 1.43 | 1.32 | 1.55 | 1.23 | 1.04 | 1.45 | 1.06 | 0.95 | 1.19 |
| Black vs. Other | 0.95 | 0.87 | 1.03 | 1.95 | 1.36 | 2.79 | 1.30 | 1.21 | 1.39 | 0.77 | 0.68 | 0.86 | 1.03 | 0.93 | 1.14 |
| Black vs. White | 0.33 | 0.32 | 0.35 | 0.80 | 0.70 | 0.92 | 0.48 | 0.47 | 0.49 | 0.26 | 0.24 | 0.27 | 0.37 | 0.35 | 0.39 |
| Hispanic vs. Other | 0.86 | 0.77 | 0.96 | 0.71 | 0.42 | 1.20 | 0.91 | 0.82 | 1.00 | 0.62 | 0.52 | 0.75 | 0.97 | 0.85 | 1.11 |
| Hispanic compared to White | 0.30 | 0.27 | 0.33 | 0.29 | 0.19 | 0.45 | 0.33 | 0.31 | 0.36 | 0.21 | 0.18 | 0.25 | 0.35 | 0.31 | 0.39 |
| Other compared to White | 0.35 | 0.32 | 0.38 | 0.41 | 0.29 | 0.59 | 0.37 | 0.34 | 0.39 | 0.34 | 0.30 | 0.37 | 0.36 | 0.33 | 0.39 |

PMAD= perinatal mood and anxiety disorders, SMI=severe mental illness, and MDP= mental disorders of pregnancy.
aRR= adjusted Relative risk, CI= Confidence Interval

### Increased risk for maternal and infant complications

For women with PMAD, we observed a higher risk of obstetric complications, including SMM (RR: 1.42, 95%CI: 1.27, 1.58), HDP (RR: 1.26, 95%CI: 1.1.19, 1.34), and CD (RR: 1.18, 95%CI: 1.15, 1.21) (Table 2). Among those with PMAD, the risk of preterm (RR: 1.40, 95%CI: 1.35, 1.45) and LBW infants (RR: 1.35, 95%CI: 1.30, 1.41) was also higher (Table 3). The incidence of LBW, HDP, and CD was higher for pregnant people with SMI. For those diagnosed with an MDP, the risk of SMM, PTB, HDP, LBW, and CD were significantly higher than deliveries without an MDP diagnosis.

**Table 3.** Adjust relative risk for maternal and infant complications for women with perinatal mental health disorder, South Carolina 1999-2017.

|  | PMAD |  |  | SMI |  |  | MDP |  |  |
| --- | --- | --- | --- | --- | --- | --- | --- | --- | --- |
| *Maternal* | **aRR** | **95%CI** |  | **aRR** | **95%CI** |  | **aRR** | **95%CI** |  |
| SMM | 1.42 | 1.27 | 1.58 | 1.35 | 0.80 | 2.25 | 1.35 | 1.21 | 1.49 |
| Cesarean | 1.18 | 1.15 | 1.21 | 1.15 | 1.02 | 1.31 | 1.25 | 1.22 | 1.28 |
| HDP | 1.26 | 1.19 | 1.34 | 1.33 | 1.02 | 1.74 | 1.23 | 1.17 | 1.30 |
| GDM | 1.20 | 1.14 | 1.27 | 1.09 | 0.82 | 1.46 | 1.25 | 1.19 | 1.32 |
| *Infant* |  |  |  |  |  |  |  |  |  |
| Preterm | 1.40 | 1.35 | 1.45 | 1.18 | 1.00 | 1.40 | 1.37 | 1.33 | 1.42 |
| LBW | 1.35 | 1.30 | 1.41 | 1.27 | 1.07 | 1.51 | 1.38 | 1.34 | 1.43 |

PMAD= perinatal mood and anxiety disorders, SMI=severe mental illness, and MDP= mental disorders of pregnancy.
aRR= Adjusted Relative risk, CI= Confidence Interval

### Increased risk for readmissions

We observed an increased risk for hospital readmissions for each maternal mental health outcome up to 45 days after the delivery event (Figure 2). For example, birthing populations were 3.9 times (95%CI: 2.73, 5.54) more likely to be readmitted if a PMAD diagnosis was present at delivery. Similarly, the risk of hospital readmission was 5-fold higher for women with an SMI at delivery and 2.6 times higher for women diagnosed with an MDP at or around the time of delivery.

**Figure 2.**
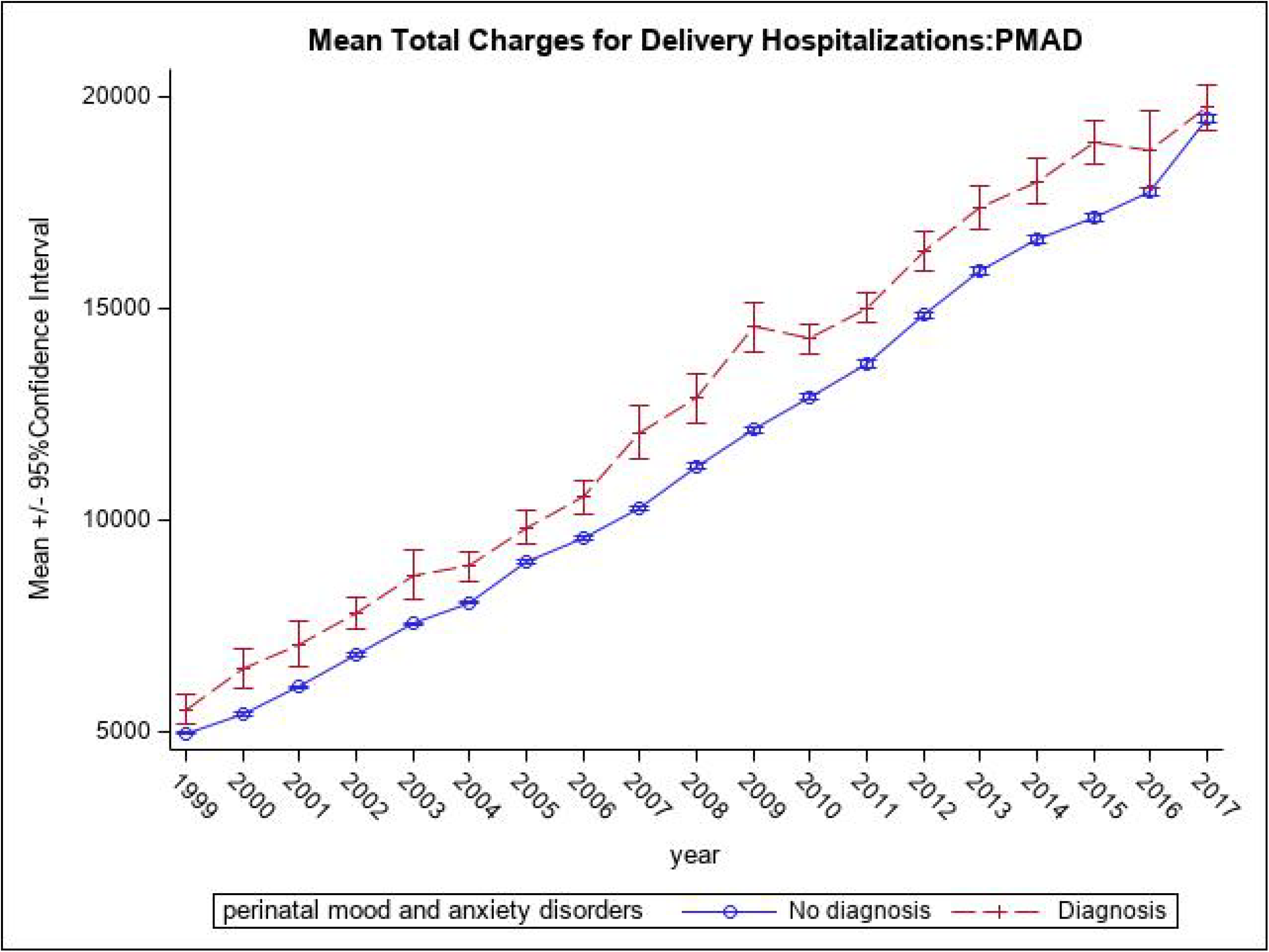
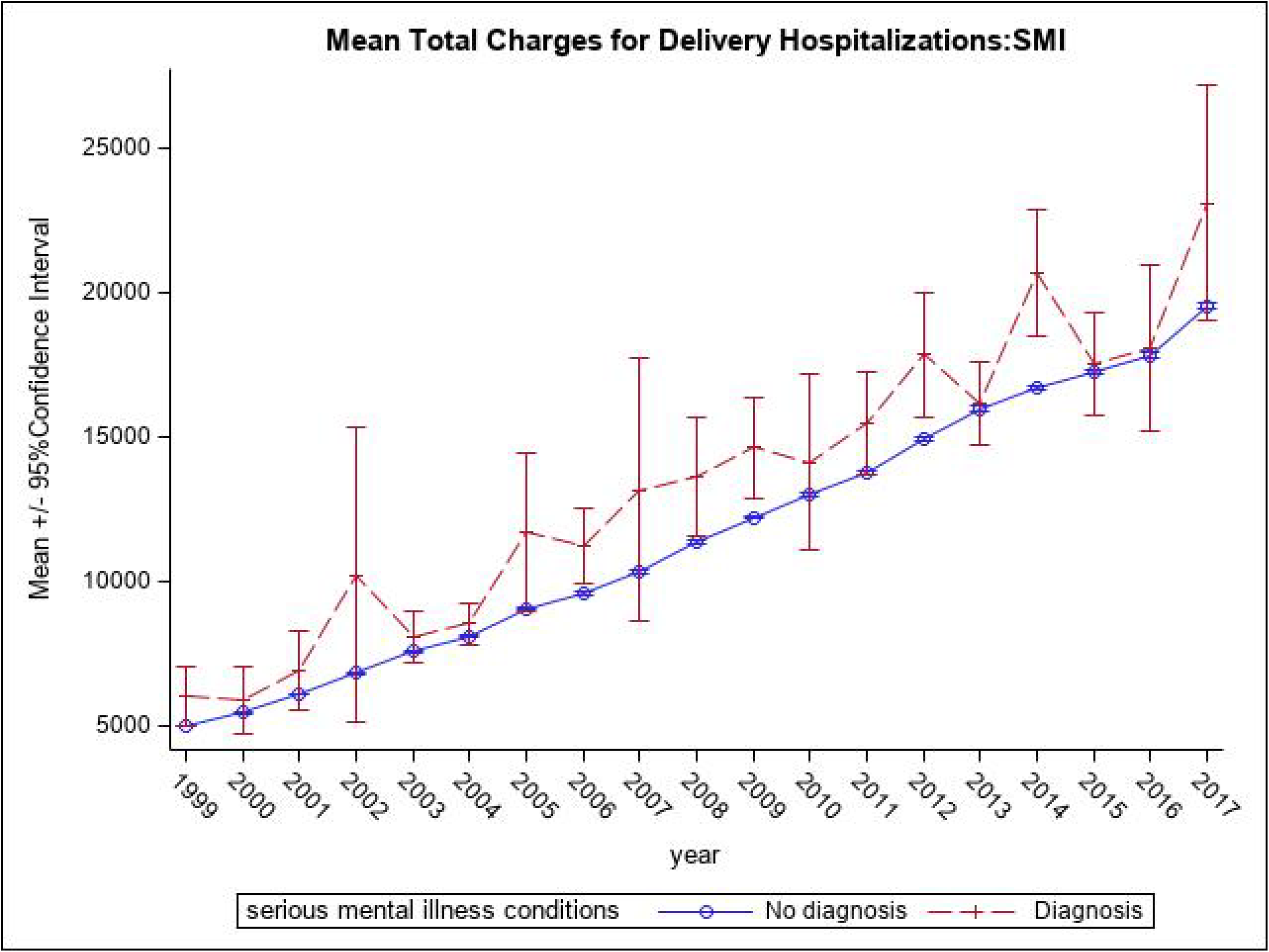
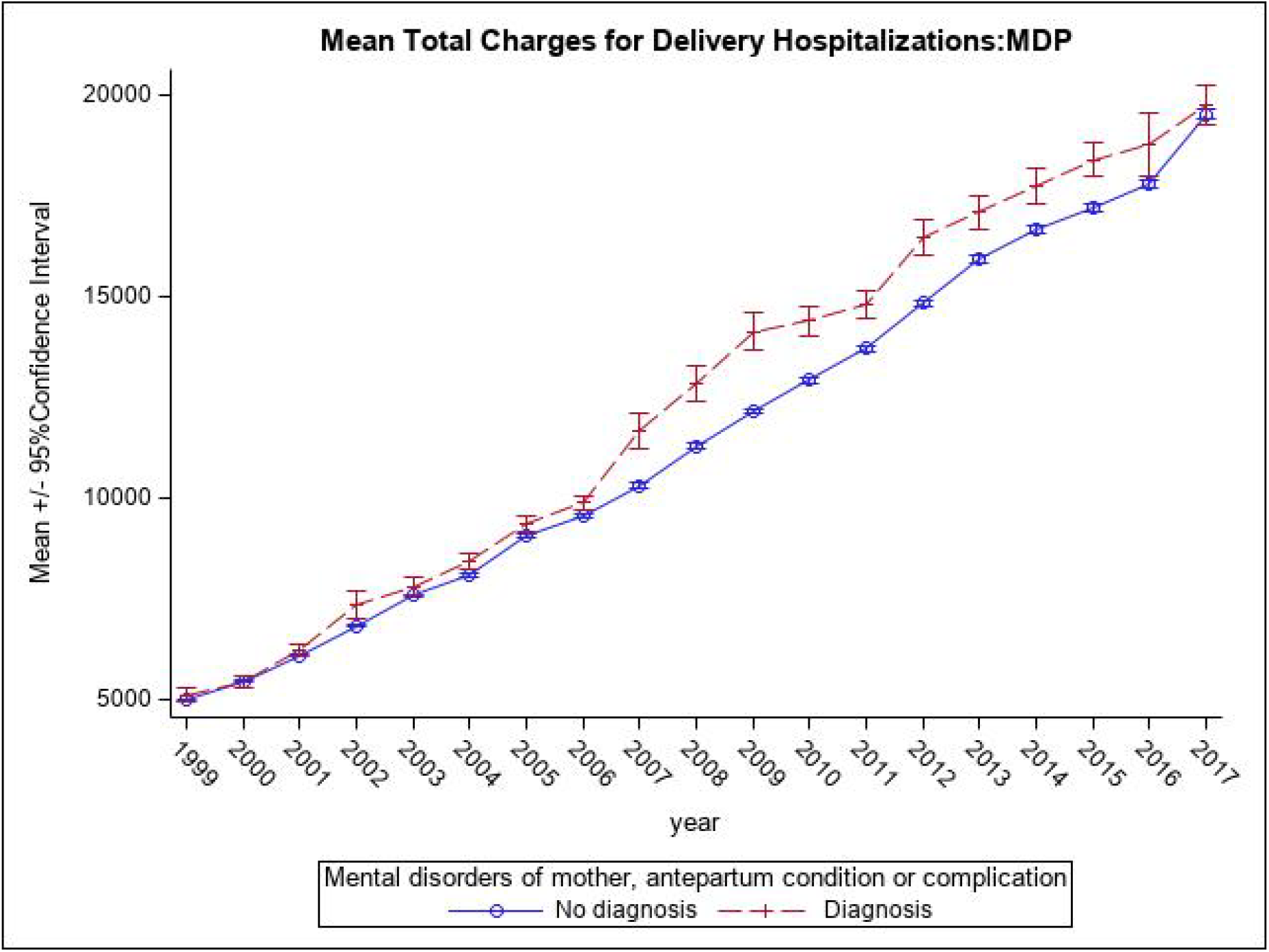
Average annual total charges per hospital delivery for a) PMAD, b) SMI, and c) MDP, South Carolina hospital deliveries 1999-2017.

### Sensitivity Analysis

Results showed elevated risks for maternal and infant complications for women with a preexisting depressive or anxiety disorder (Table S3). For pregnant persons with a depression or anxiety diagnosis during or around the time of delivery, there was excess risk for each maternal and infant complication. However, higher risk for SMM, CD, HDP, GDM, PTB, or LBW complications were detected among women with a perinatal anxiety disorder compared to women without anxiety.

### Higher cost of Deliveries with Mental Health Conditions

The average cost of a hospital delivery associated with a PMAD ($4,029 more per delivery), SMI ($1,956 more per delivery), or MDP diagnosis ($1,968 more per delivery) was significantly higher than deliveries not related to a mental health outcome (Table 4), and these costs have been on the rise (Figure 3).

**Table 4.** Descriptive statistics on total charges per mental health-related delivery, South Carolina 1999-2017.

| *PMAD* | Total deliveries | Mean Difference | Mean | 95% CI mean |  | Median | IQR |  | P-value |
| --- | --- | --- | --- | --- | --- | --- | --- | --- | --- |
| No | 893337 |  | 11588 | 11571 | 11606 | 9442 | 6304 | 14460 | <.001 |
| Yes | 24199 | 4029 | 15617 | 15457 | 15777 | 13113 | 9052 | 19251 |  |
| *SMI* |  |  |  |  |  |  |  |  |  |
| No | 916280 |  | 11692 | 11674 | 11710 | 9525 | 6353 | 14606 | <.001 |
| Yes | 1256 | 1956 | 13648 | 13015 | 14280 | 11093 | 7377 | 16606 |  |
| *MDP* |  |  |  |  |  |  |  |  |  |
| No | 877780 |  | 11609 | 11591 | 11627 | 9461 | 6313 | 14503 | <.001 |
| Yes | 39756 | 1968 | 13577 | 13466 | 13688 | 11068 | 7445 | 16962 |  |

PMAD= perinatal mood and anxiety disorders, SMI=severe mental illness, and MDP= mental disorders of pregnancy. CI= Confidence Interval, IQR=Interquartile range

**Figure 3.**
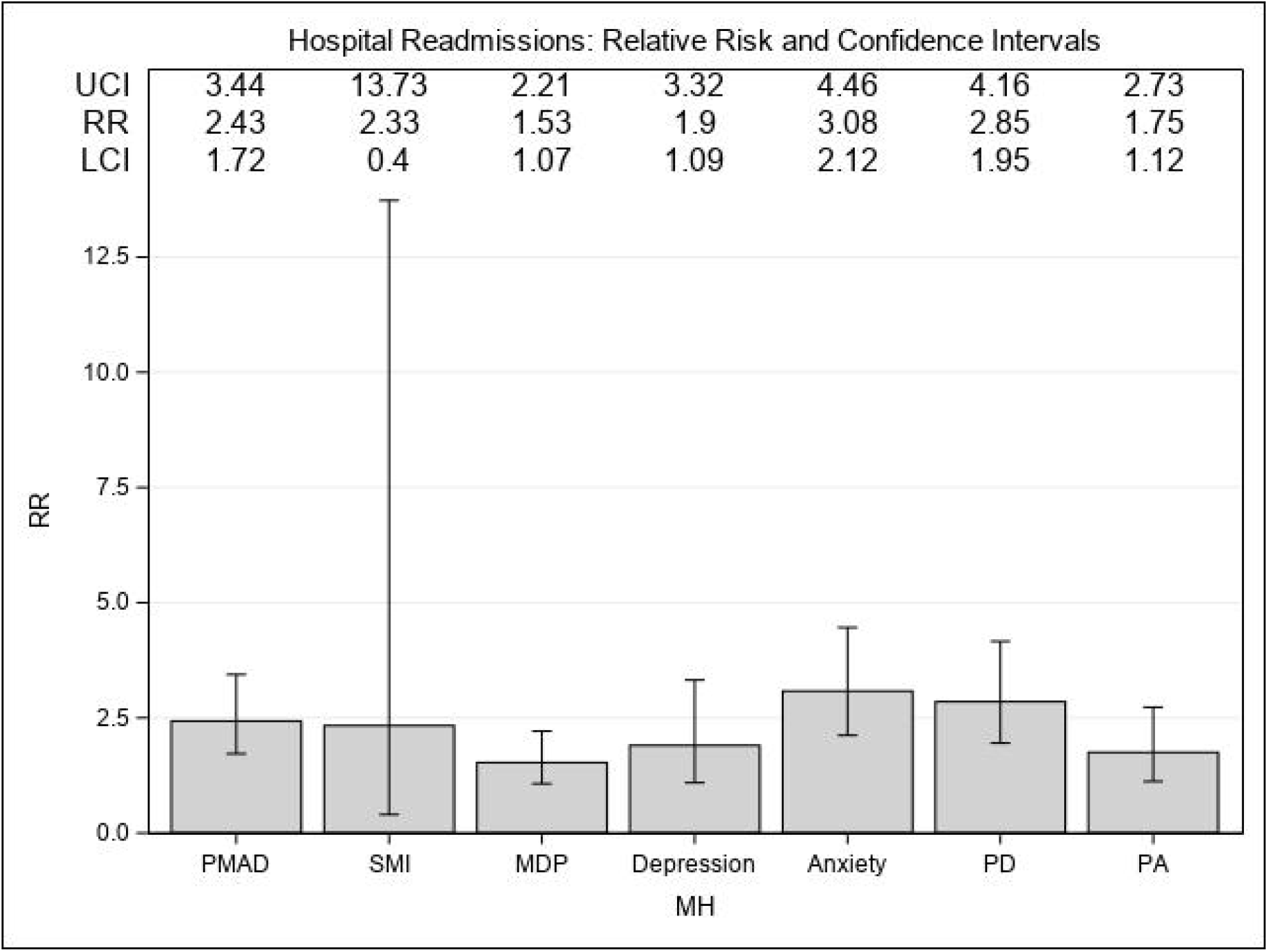
The relative risk (RR) and associated 95% confidence interval (CI) for Perinatal mood and anxiety disorders (PMAD), Severe maternal illness (SMI), Preexisting depression (PD), and Preexisting anxiety (PA), South Carolina hospital readmissions 1999-2017.

## DISCUSSION

This study is the first to examine maternal morbidity associated with perinatal mental health disorders in the Southeastern U.S.. Similar to other studies, our results demonstrated the escalating burden of PMAD and MDP for pregnant populations over time with important consequences related to maternal and infant morbidity (12). SMI was the most important contributor to hospital readmissions in the postpartum period. A PMAD or MDP diagnosis during or around the time of delivery was associated with an increased risk of adverse maternal complications, such as severe maternal morbidity, hypertensive disorders of pregnancy, cesarean delivery, gestational diabetes, and adverse infant outcomes like preterm delivery and low birth weight. SMI was an important maternal risk factor for cesarean delivery and hypertensive disorders of pregnancy in pregnant populations and infants with low birth weight. Compared to mood disorders, perinatal anxiety disorders contributed the most to maternal and infant morbidity. Maternal factors like Medicaid coverage, residence in an urban area, substance abuse, obesity, and multiparity, were significantly associated with increased risk for each maternal mental disorder. Lastly, PMAD, SMI, and MDP were associated with significantly higher delivery costs revealing the cost burden of mental health morbidity for families in the South.

Pregnancy is both a vulnerable and life-altering time, in which a woman undergoes several physical, social, and psychological adjustments. Often a woman’s experience of obstetric complications during pregnancy, like the diagnosis of gestational hypertension or diabetes, or the unanticipated cascade of changes for either mom or baby that result in an emergency cesarean section can be followed by mental health sequelae in the perinatal and postpartum periods. For example, compared to vaginal deliveries, persons who experienced a CD were much more likely to be diagnosed with depression, anxiety, obsessive compulsive, or somatic symptom disorders in the three months (30) and six months (31) after delivery. In more severe cases, CD has been associated with the most severe psychiatric disorders, including suicidal behavior and attempts, up to one year after delivery (32). Maternal mental health and well-being should be a focal concern during the obstetric encounters in the year following childbirth, especially if women present with a history of mental illness or experience an obstetric or infant complication (33). We observed a strong association between SMM and a PMAD or MDP diagnosis, whereby anxiety disorders were the largest contributor to SMM risk. A recent U.S.-based population health study documented that birthing populations with a mental health disorder were 1.5 times more likely to experience SMM compared to those without (16). Another Swedish cohort study showed that persons recovering from an SMM delivery were much more likely to receive treatment for a psychiatric disorder in the year post-delivery (17).

Our study revealed an excess risk of GDM in the presence of PMAD or MDP diagnosis. The linkage between GDM and a greater risk of prenatal or postpartum mental health disorders is well supported in the literature, particularly among racially and ethnically diverse populations (34, 35). In fact, women diagnosed with GDM were two-fold more likely to experience pregnancy or postpartum-related depression than women without a GDM diagnosis (35). Results from a recent systematic review suggest a strong connection between psychosocial well-being and dietary and lifestyle behaviors (e.g., physical fitness) for pregnant women encountering a GDM diagnosis (36). More interventional research, especially during pregnancy, should couple psychological and social support with lifestyle interventions to reduce the short-term psychological risk and potentially safeguard against GMD-related cardiovascular and metabolic disorders later in life (35).

Similar to results from our study, prenatal anxiety and depressive disorders have also been shown to significantly increase a person’s risk for a HDP diagnosis, and in some instances result in a shorter gestational period (37). Results from a prospective Mexican cohort showed that anxiety, sleep or social dysfunction, and acute somatic symptoms at 20 weeks of gestation were associated with a higher risk of developing HDP (38). Preexisting depression or anxiety disorder in the one year before conception has been associated with chronic hypertension and preeclampsia during pregnancy (39), and limited research has shown that active-duty women with a PTSD diagnosis before conception were more likely to experience an HDP (40).

Birthing populations who experience maternal psychological distress, like pregnancy-related anxiety or mood disorders, have an elevated risk of PTB (41). Similar to other population-based cohort studies, we observed SMI (includes the diagnosis of a bipolar disorder) and depressive disorders were associated with excess risk for PTB (42). Some research has shown that PTB coupled with other maternal somatization disorders may result in a higher risk for mood and anxiety disorders for postpartum women (43) and children (41). One mechanism linking maternal distress with adverse infant health outcomes is parental confidence. One study showed that following a PTB involving a CD, a reduction in confidence levels was observed. Still, maternal distress has also been observed following the delivery of full-term babies (44). The persistence of maternal distress during pregnancy and the postpartum period is a leading risk factor for impaired mother-child attachment, infant developmental delays, and childhood and later in life health risks (18, 45). Reducing maternal distress should be one central focus of clinic and home-based interventions. New health policies are needed to ensure prolonged support for women from socially disadvantaged backgrounds or those whose infant experienced neonatal complications.

For our sample, we observed a higher risk of LBW for women with SMI. Very few studies have looked at the association between SMI and LBW (46). We found only one study that showed an episode of SMI during pregnancy was associated with nearly a three-fold risk of having a LBW infant (47). Compared to women with no history of mental illness, a retrospective birth cohort study examining women with schizophrenia or major affective disorders (i.e., proxy for SMI) in Australia showed no association with LBW (48), while a Taiwan-based study showed higher risk of LBW for women with bipolar disorder (49).

People with a mental health condition at or around the time of delivery also incur higher medical costs per delivery. One national study showed the mean costs are $458 higher per delivery discharge event (16). We observed an even higher mean cost per delivery ranging from $1968 for an MDP event to $4029 per delivery associated with a PMAD diagnosis. Another study found non-Latina Black women experience both higher rates of prenatal depressive symptoms and use significantly lower volume of postpartum counseling services and medications than non-Latina White women (50).

Prior research has shown that roughly two women in 1,000 are at elevated risk for an incident or exacerbated episode of severe mental illness in the early weeks of the postnatal period (51-53). Our study showed that women with PMAD or SMI were more than two-fold more likely to be readmitted to the hospital. Women in our sample who were diagnosed with a perinatal anxiety disorder were three times as likely to be readmitted for inpatient care in the 45 days following delivery. Results also demonstrated increased risk for hospital readmission in the postnatal period among birthing populations diagnosed with MDP (i.e., 150% increase), depressive disorders (i.e.,190% increase), and who had a prior history of depression (i.e., 285% increase) or anxiety (i.e.,175% increase). A previous study found that the risk of psychiatric readmission for women was 10 to 19 days post-delivery; whereby a preexisting diagnosis of bipolar affective disorder was the strongest risk factor (54).

While white women were the most at risk in our study sample, results may indicate maternal mental disorders are an under-diagnosed pregnancy risk in Black birthing populations. Black pregnant populations in our sample were much less likely to be diagnosed with a prenatal or perinatal mental disorder compared to whites. Because we relied on hospital administrative or billing data, we can only speculate why this may be the case. Racialized pregnancy stigma has surfaced as one potential explanation for why Black mothers may experience discriminatory or racialized stigmatizing that includes reduced access to quality and timely health services, resources, and support. One study using a small sample of pregnant women in Connecticut found that Black women repeatedly encountered racialized pregnancy stigma in health care, a persistent source of maternal stress (55). Further, this issue around the experience of racism and pregnancy is specific to the U.S. and racism-related stress may have adverse consequences for both mom and baby, including declines in mental health status (56). Findings from a prospective cohort study on pregnant women in Boston, Massachusetts, U.S. showed that the self-reported prevalence of personally experienced and the group experienced racism was 410% and 780% higher, respectively, for U.S.-born compared to foreign-born Black pregnant populations (57).

Elevated depression symptoms have been associated with perceived racial discrimination in Black pregnant women in the U.S. (58). Moving beyond individual experience of racism, research has also shown that exposure to structural racism (racialized housing, education, employment, and health care patterns and practices that reinforce discriminatory beliefs, values, and distribution of resources) during pregnancy may adversely effect maternal and infant health outcomes for Black women(59). A qualitative study showed that Black pregnant and postpartum women residing in racially and economically deprived neighborhoods (i.e., a proxy for structural racism) in Oakland, California, U.S. were 54.5% more likely to experience racial discrimination while receiving medical care (60). In addition to the experience of racism among pregnant women, past childhood exposure to racism and environmental trauma has been associated with an increased risk for perinatal mental illness (61).

### Strengths and Limitations

To our knowledge, this is one of the first longitudinal studies to examine maternal mental health complications during pregnancy and the perinatal period in a Southern state characterized by poor reproductive and infant health outcomes. We were able to examine the incidence of maternal mental disorders over time for a lengthy study period (i.e., 1999-2017) and for women with multiple delivery events. Further, we disaggregated PMAD into contributing depressive or anxiety-related disorders, and we also looked at the influence of a preexisting diagnosis for anxiety or depression.

Our results are subject to a few limitations. We focused our analysis on one state in the Southern U.S., but more research should include data from other states in the South. We relied on hospital administrative data used for service billing and were unable to obtain detailed data on maternal income, marital status, and family structure. Data on paternal factors, like mental health disorders or treatment need, were also unavailable. Pregnant populations with preexisting mental health conditions may have discontinued prenatal prescription drug use for fear of harming the infant. Discontinuation is particularly common among women with bipolar disorders, and our analysis did not consider discontinuation or the potential complications introduced by stopping medication use while pregnant. Lastly, our data on maternal mental health disorders are subject to bias from coding and diagnostic errors, but prior research has shown higher reliability for well-defined disorders such as psychotic disorders (62).

### Policy and/or clinical implications

Following a life course approach, there is significant evidence to suggest that the experience of poverty, poor physical health, interpersonal violence, and other types of social disadvantage accumulate over the life span to influence mental health, particularly during the sensitive period occurring around the time of childbirth (63). Additional research on innovative strategies and well-timed clinical and public health interventions during the preconception, prenatal, and postpartum periods is needed to reduce the burden of maternal mental health for women and their families. As the pandemic continues, a closer examination of changes in maternal mental health and the associated reproductive and neonatal consequences will inform the timing and components of interventions for a future health crisis.

### Conclusion

Our findings highlight the significant contribution of maternal mental health during and around the time of pregnancy to reproductive and infant health complications for a Southern birth cohort. Perinatal mood and anxiety disorders and mental disorders debuting during pregnancy were positively associated with increased risk of severe maternal morbidity, hypertensive disorders of pregnancy, and gestational diabetes, as well as increased neonatal risks arising from a preterm birth or low birth weight at delivery. Birthing populations diagnosed with severe maternal mental illness were at greater risk for a cesarean delivery or low birth weight infant. When disaggregating mood and anxiety disorders, results showed that anxiety was the most significant predictor of maternal and infant health complications. Perinatal mental disorders are a commonly recognized maternal phenomenon during the reproductive years. Still, more evidence-based interventions at the level of the individual and population-based public health interventions are needed to reduce the health, psychological, and socio-economic burden of these conditions on women and their families.

## Data Availability

All data produced in the present work are contained in the manuscript

## Acknowledgement

We acknowledge Chris Finney and Sarah Crawford at the SC Revenue and Fiscal Affairs Office Health and Demographics division for all of their data support.

## Authorship contribution statement

JR conceptualized the design and methodological approach, conducted the analysis and drafted the paper; KR participated in the drafting of the paper; MR participated in the design and the drafting of the paper; and MS participated in the design and the drafting of the paper.

## Notes

### Competing Interest Statement

The authors have declared no competing interest.

### Funding Statement

This study did not receive any funding

### Author Declarations

Ethics committee/IRB of NC State University gave ethical approval for this work Ethics committee/IRB of Appalachian State University gave ethical approval for this work

