## Supplemental Materials for "Association between perinatal mental health and reproductive and neonatal complications: a retrospective birth cohort study"

**Supplemental Tables**

Supplemental Table 1. List of ICD-9 diagnosis codes used to define maternal mental health outcomes, reproductive complications, and some covariates.

| ***Maternal mental health*** | |  |
| --- | --- | --- |
| PMAD | Depression | *Major depressive affective disorder* 296.20, 296.21, 296.22, 296.23, 296.24, 296.25, 296.26, 296.30, 296.31, 296.32, 296.33, 296.34, 296.35, 296.36. *Dysthymic disorder* 300.4. *Depressive disorder,* not elsewhere classified 311 |
|  | Anxiety | *Anxiety disorder* 293.84, 300.00, 300.01, 300.02, 300.09, 300.10. *Phobia* 300.20, 300.21, 300.22, 300.23, 300.29. *Obsessive-compulsive disorder* 300.3. *Neurasthenia and other somatoform disorders* 300.5, 300.89, 300.9. *Acute stress reaction* 308.0, 308.1, 308.2, 308.3, 308.4, 308.9. *Posttraumatic Stress Disorder* 309.81. *Overanxious disorder* 313.0, 313.1, 313.21, 313.22, 313.30, 313.82, 313.83. |
| SMI | Bipolar disorder | *Bipolar I disorder* 296.00, 296.01, 296.02, 296.03, 296.04, 296.05, 296.06, 296.40, 296.41, 296.42, 296.43, 296.44, 296.45, 296.46, 296.50, 296.51, 296.52, 296.53, 296.54, 296.55, 296.56, 296.60, 296.61, 296.62, 296.63, 296.64, 296.65, 296.66, 296.7. Manic disorder 296.10, 296.11, 296.12, 296.13, 296.14, 296.15, 296.16. *Other and unspecified bipolar disorders* 296.80, 296.81, 296.82, 296.89. *Other and unspecified episodic mood disorder* 296.90, 296.99. |
|  | Psychotic Disorders | *Psychotic disorder* 293.81, 293.82. *Schizophrenia* 295.00, 295.01, 295.02, 295.03, 295.04, 295.05, 295.10, 295.11, 295.12, 295.13, 295.14, 295.15, 295.10, 295.11, 295.12, 295.13, 295.14, 295.15, 295.20, 295.21, 295.22, 295.23, 295.24, 295.25, 295.30, 295.31, 295.32, 295.33, 295.34, 295.35, 295.40, 295.41, 295.42, 295.43, 295.44, 295.45, 295.50, 295.51, 295.52, 295.53, 295.54, 295.55, 295.60, 295.61, 295.62, 295.63, 295.64, 295.65, 295.70, 295.71, 295.72295.73, 295.74, 295.75, 295.80, 295.81, 295.82, 295.83, 295.84, 295.85, 295.90, 295.91, 295.92, 295.93, 295.94, 295.95. *Paranoid state* 297.0297.1297.2297.3297.8297.9. *Depressive* *psychosis* 298.0, 298.1, 298.2, 298.3, 298.4, 298.8, 298.9. |
| MDP | Mental disorders complicating pregnancy childbirth or the puerperium | 648.4 |
| ***Maternal Complications*** | |  |
| SMM | | 1. Acute myocardial infarction DX 410.xx; 2. Aneurysm DX 441.xx; 3. Acute renal failure DX 584.5, 584.6, 584.7, 584.8, 584.9, 669.3x; 4. Adult respiratory distress syndrome DX 518.5x, 518.81 518.82 518.84, 799.1; 5. Amniotic fluid embolism DX 673.1x; 6. Cardiac arrest/ventricular fibrillation DX 427.41, 427.42, 427.5; 7. Conversion of cardiac rhythm PR 99.6x; 8. Disseminated intravascular coagulation DX 286.6, 286.9, 666.3x; 9. Eclampsia DX 642.6x; 10. Heart failure/arrest during surgery or procedure DX 997.1; 11. Puerperal cerebrovascular disorders DX 430.xx, 431.xx, 432.xx, 433.xx, 434.xx, 436xx, 437.xx, 671.5x, 674.0x, 997.02; 12. Pulmonary edema / Acute heart failure DX 518.4, 428.1, 428.0, 428.21, 428.23, 428.31, 428.33, 428.41, 428.43; 13. Severe anesthesia complications DX 668.0x, 668.1x, 668.2x; 14. Sepsis DX 038.xx, 995.91, 995.92, 670.2x; 15. Shock DX 669.1x, 785.5x, 995.0, 995.4, 998.0x; 16. Sickle cell disease with crisis DX 282.42, 282.62, 282.64, 282.6; 17. Air and thrombotic embolism DX 415.1x, 673.0x, 673.2x 673.3x, 673.8x; 18. Blood products transfusion PR 99.0x; 19. Hysterectomy PR 68.3x-68.9x; 20. Temporary tracheostomy PR 31.1; 21.Ventilation PR 93.90, 96.01, 96.02, 96.03, 96.05 |
| GDM | | 648.8 |
| CD | | Diagnosis codes:765, 766; Procedure codes: 740, 741, 742, 744, 749.9; Diagnosis Related Group (DRG) type: '765', '766' |
| HDP | | 642.3, 642.4,642.5,642.6,642.7 |
| ***Covariates*** | |  |
| Substance Abuse | | 303.93, 305.00, 305.01, 305.02, 305.03, 357.5, 425.5, 535.30, 535.31, 571.0, 571.1, 571.2, 571.3, E86.00, 304.40, 304.41, 304.42, 304.43, 305.70, 305.71, 305.72, 305.73, 304.30, 304.31, 304.32, 304.33, 305.20, 305.21, 305.22, 305.23, 304.20, 304.21, 304.22, 304.23, 305.60,305.61, 305.62, 305.63, 968.5, E938.5, 292.11, 292.12, 292.2, 292.81, 292.82, 292.83, 292.84, 292.85, 292.89,292.9, 304.50, 304.51, 304.52, 304.53, 304.53, 305.30, 305.31, 305.32, 305.33, 969.6, E854.1, E939.6, 304.00,304.01, 304.02, 304.03, 304.70, 304.71, 304.72, 304.73, 305.50, 305.51,305.52, 305.53, 965.00, 965.01,965.02, 965.09, E85.00, E935.0, 304.10, 304.11, 304.12, 304.13, 305.40, 305.41, 305.42, 305.43, 304.60, 304.61, 304.62,304.63, 304.80, 304.81, 304.82, 304.83, 304.90, 304.91, 304.92, 304.93, 305.90, 305.91, 305.92, 305.93, 648.30, 648.31, 648.32, 648.33, 648.34, V654.2 |

PMAD= Perinatal mood and anxiety disorders, SMI=Serious mental illness conditions, MDP=Mental disorders complicating pregnancy childbirth or the puerperium, SMM=Severe maternal morbidity, GDM= Gestational diabetes mellitus, CD= Cesarean delivery, HDP= Hypertensive disorders of pregnancy.

Table S2. Prevalence of maternal mental health associated hospital delivery and hospital readmission visits, South Carolina 1999-2017.

|  | PMAD | SMI | MDP | Depression | Anxiety | PD | PD |
| --- | --- | --- | --- | --- | --- | --- | --- |
| Hospital Deliveries | n (%) | n (%) | n (%) | n (%) | n (%) | n (%) | n (%) |
| Yes | 24201 (2.7) | 1230 (0.13) | 35435 (3.9) | 15607 (1.7) | 11425 (1.3) | 19833 (2.2) | 13602 (1.5) |
| No | 889129 (97.4) | 912100 (99.9) | 877895 (96.1) | 897723 (98.3) | 901905 (98.8) | 893497 (97.8) | 899728 (98.5) |
| Readmission |  |  |  |  |  |  |  |
| Yes | 1021 (4.0) | 101 (7.4) | 2026 (4.9) | 719 (3.2) | 500 (2.2) | NA | NA |
| No | 24207 (96.0) | 1256 (92.6) | 39,773 (95.2) | 15599 (1.7) | 11444 (1.5) | NA | NA |

Table S3. Adjusted relative risk for maternal and infant complications for women with depression, anxiety, or preexisting depression or anxiety, South Carolina 1999-2017.

|  | Depression | | | Anxiety | | | PD | | | PA | | |
| --- | --- | --- | --- | --- | --- | --- | --- | --- | --- | --- | --- | --- |
| *Maternal* | **aRR** | **95%CI** | | **aRR** | **95%CI** | | **aRR** | **95%CI** | | **aRR** | **95%CI** | |
| SMM | 1.35 | 1.17 | 1.56 | 1.66 | 1.43 | 1.92 | 1.33 | 1.17 | 1.51 | 1.67 | 1.45 | 1.91 |
| Cesarean | 1.11 | 1.08 | 1.15 | 1.24 | 1.19 | 1.29 | 1.13 | 1.10 | 1.17 | 1.24 | 1.19 | 1.28 |
| HDP | 1.18 | 1.10 | 1.28 | 1.34 | 1.24 | 1.46 | 1.18 | 1.10 | 1.26 | 1.35 | 1.25 | 1.45 |
| GDM | 1.18 | 1.10 | 1.27 | 1.24 | 1.15 | 1.35 | 1.19 | 1.11 | 1.26 | 1.22 | 1.13 | 1.31 |
| *Infant* |  |  |  |  |  |  |  |  |  |  |  |  |
| Preterm | 1.37 | 1.31 | 1.44 | 1.45 | 1.37 | 1.53 | 1.38 | 1.32 | 1.44 | 1.46 | 1.39 | 1.54 |
| LBW | 1.34 | 1.27 | 1.41 | 1.38 | 1.30 | 1.47 | 1.36 | 1.30 | 1.43 | 1.40 | 1.33 | 1.49 |
| PD=Preexisting depression, PA=Preexisting Anxiety, SMM= Severe Maternal Morbidity, HDP= Hypertensive Disorders of Pregnancy, GDM= Gestational Diabetes Mellitus, LBW=Low Birth Weight | | | | | | | | | | | | |
| RR= adjusted Relative risk, CI= 95% Confidence Interval | | | | | | | | | | | | |
